# Bibliometric analysis of published articles on perinatal anxiety from 1920-2020

**DOI:** 10.1101/2023.09.29.23296340

**Authors:** Justine Dol, Marsha Campbell-Yeo, Patricia Leahy-Warren, Chloe Hambly LaPointe, Cindy-Lee Dennis

## Abstract

**Introduction:** Trends and gaps in perinatal anxiety research remain unknown. The objective of this bibliometric review was to analyze the characteristics and trends in published research on perinatal anxiety to inform future research.

**Methods:** All published literature in Web of Science on perinatal anxiety from January 1, 1920 to December 31, 2020 were screened by two reviewers. VOSViewer was utilized to visualize linkages between publications. Bibliometric data were extracted from abstracts.

**Results:** The search strategy identified 4,561 publications. After screening, 2,203 publications related to perinatal anxiety were used for the visualization analysis. For the bibliometric data, 1,534 publications had perinatal anxiety as a primary focus. There were 7,910 different authors, over half named only once (55.5%), from 63 countries. 495 journals were identified, with over half (56.0%) publishing only one article. Most articles were published between 2011-2020 (75.9%). In terms of perinatal timing, over half (54.2%) published on antenatal anxiety. Only 6.0% of studies reported on perinatal anxiety in fathers and 56.5% reported on postpartum depression.

**Limitations:** Web of Science was solely used, and manual screening of each publication was required.

**Conclusion:** This bibliometric analysis found: (1) perinatal is a growing field of research, with publications increasing over time; (2) there is variation in authors and journals; (3) over half of the publications focus on antenatal anxiety; (4) paternal anxiety is understudied; and (5) only 6% of publications came from low and lower-middle income countries. Gaps related to maternal postnatal, and all paternal perinatal anxiety exist.

## INTRODUCTION

There has been significant growth in the literature on anxiety across the perinatal period with major variations in the conceptual definition and measurement of it as a mental health outcome (Meades and Ayers, 2011; Sinesi et al., 2019; Ukatu et al., 2018) (Leach et al., 2017; Norhayati et al., 2015; Wee et al., 2011). While anxiety is the most common mental illnesses diagnosed each year worldwide (Ströhle et al., 2018), in the perinatal population it can emerge during pregnancy or up to one year post-birth and is shaped by fear and worry, particularly related to pregnancy outcomes or newborn care. It can present as generalized anxiety, panic disorders, obsessive-compulsive disorder, or post-traumatic stress disorder (Ali, 2018) and effects up to 39% of women (Fairbrother et al., 2016; Leach et al., 2017) and 18% of fathers (Leach et al., 2016; Leiferman et al., 2021).

To focus future research and identify gaps in perinatal anxiety, it is important to explore patterns and trends in the literature. While a few reviews on anxiety across the perinatal period exist, none have used a bibliometric approach to map the evolution of the published literature to characterize research outputs, distribution, and relationships. The purpose of this bibliometric analysis is to examine the characteristics and trends in publications on anxiety across the perinatal period from January 1, 1920 to December 31, 2020. The specific aims are to (1) identify, analyze, and visualize research publications on perinatal anxiety, and (2) explore outcomes and co-relations such as author, country, institution, and year in order to present a comprehensive overview of perinatal anxiety publications over the last century.

## METHODS

This study follows the standard bibliometric approach (Donthu et al., 2021; Gauthier, 1998; Linnenluecke et al., 2020), using the following steps: (1) define the search criteria, keywords, and time periods; (2) selection of Web of Science database; (3) adjustment and refinement of research criteria; (4) full export of result; (5) analysis of the information and discussion of the results (Ruiz-Real et al., 2018, p. 2). A preliminary search of MEDLINE, the Cochrane Database of Systematic Reviews and PROSPERO was conducted and no current or underway systematic reviews, scoping reviews, or bibliometric analysis on the topic were identified. This study follows a predefined protocol (Dol et al., 2021).

### Search Strategy

The search strategy was conducted in Web of Science using the Web of Science Core Collection. The search strategy that was developed with a health science librarian and authors was: TS=((pregnan* OR perinatal* OR antenatal* OR postnatal* OR peripartum OR postpartum OR “post partum” OR “new mother*” OR “new father*” OR “new parent*” OR (after NEAR/2 birth*) OR paternal* OR maternal*) NEAR/3 (anxiet* OR anxious*)). The search was limited to dates between 1920 and 2020.

### Inclusion and Exclusion Criteria

All publications included were either a review (systematic or otherwise) or article (any design) published between January 1, 1920 to December 31, 2020 in any language. Conference abstracts, books/book reviews, theses, and non-peer reviewed material as defined by the database were excluded. For the purpose of this study, the term ‘publication’ will be used to refer to all identified literature.

After a preliminary review of the included publications, it was discovered that the search strategy pulled a significant number of studies not related to perinatal anxiety. Therefore, it was decided that it was necessary to screen all publications to ensure relevancy to perinatal anxiety, resulting in a deviation from protocol which did not include a screening step. All studies were uploaded into Covidence and were independently screened by the first author (JD) and fourth author (CLH) by checking title and/or abstract to determine if it was relevant to perinatal anxiety, with disagreements resolved by a third author. Publications included in the visualization analysis must have mentioned perinatal anxiety as a primary, secondary, or covariate outcome and reported on the related outcomes in the findings and methods. Publications included in the bibliometric data must have had perinatal anxiety as a primary focus. Publications were excluded from both analysis if they reported on (1) separation anxiety, (2) maternal or paternal anxiety beyond one-year postpartum, (3) other perinatal mental health concerns (e.g., stress, depression), or (4) animal-based studies.

### Data Analysis

#### Bibliometric Data

After reviewing the identified articles from the search, it was determined necessary to screen publications for relevancy to perinatal anxiety. As a result, Web of Science was no longer able to be used for the bibliometric data analysis. Instead, bibliometric information on number of publications, year of publication, authors, journals, and countries were cleaned and calculated in Excel. Additionally, a data extraction survey was conducted on the abstracts of all relevant publications. The data extraction form was used to first determine if perinatal anxiety was a primary focus of the publication or not. If yes, data were extracted on the following outcomes using only the abstract: country where study was conducted, type of study, timing of anxiety, if fathers were included, if postpartum depression was also reported on, and type of measurement tool used. Double data verification was completed on 7% (n=160) of publications with an 88.2% agreement rate.

#### Visualization Mapping

VOSViewer was utilized to visualize the networks of linkages between publications, including bibliometric networks of co-authorship, co-occurrence, and co-citation, as well as co-occurrence between keywords (excluding non-relevant or generic terms) (van Eck and Waltman, 2020). Publications that were deemed not related to perinatal anxiety at the title/abstract screening phase were removed from the data files used for analysis. Analyses and visualizations were conducted for co-authorship, keyword co-occurrence, citation, bibliographic coupling, and co-citation. Additionally, co-occurrence of keywords was conducted based on text data using title and abstract data. The size of a node, or relationship, is indicative of frequency, with larger nodes representing a higher number of items identified, with the line between two nodes indicating connection between items. Nodes of the same color belong to a cluster or grouping of similar items.

## Results

The search strategy initially identified 4,561 publications, which were uploaded to Covidence for screening. Three publications were removed as a duplicate and 2,355 were removed as ineligible, leaving 2,203 publications included for the visualization analysis. Of these, 647 were deemed not to have perinatal anxiety as a primary focus (e.g., reported as a covariate, secondary outcome, etc.) and 12 were considered unclear with no abstract available, thus were excluded from the bibliometric analysis, leaving 1,534 publications in the bibliometric analysis.

### Bibliometric Data

Among the 1,534 publications, there were 7,910 different authors, most listed only once (n=4407, 55.7%) or twice (n=690, 8.7%). The author with the highest number of publications was Glover V at 35 (1.05%). For first listed author, there were 1,214 different authors, meaning that only 20.8% of publications has a first listed author more than once. Matthey,S, Reck,C, and Uguz,F were tied with the most first-authored publications at eight each. Publications had a mean and median of 5 authors, with a range between 1 and 111.

In terms of study design, most were quantitative (79.9%), followed by reviews (8.4%), questionnaire development or validation (4.4%), qualitative (1.5%), and mixed methods (1.2%). Nine percent were other study types including protocols, case studies, and discussion papers. Over half (54.2%) published on antenatal anxiety only, followed by 22.7% reporting on both antenatal and postpartum anxiety (i.e., perinatal), 21.1% on postpartum anxiety only, 1.7% on intrapartum anxiety only, and 0.3% having unclear timing. Only 6.0% of studies reported on anxiety in fathers. Across all studies, 56.5% reported on postpartum depression. Of the 1,225 quantitative studies, 62.3% reported the type of measurement tool used in the abstract. Of the measures used to assess perinatal anxiety among quantitative studies, State-Trait Anxiety Inventory (Spielberger et al., 1983) was reported inmost publications (52.7%), followed by clinical interviews (9.0%), and the Hospital Anxiety Depression scale (Zigmond and Snaith, 1983) (6.9%).

There were 63 countries identified based on corresponding author, with the United States being the highest publishing country at 20.5% of publications, followed by United Kingdom of Great Britain and Northern Ireland (9.2%), Australia (8.9%), and Canada (8.1%). According to the income classifications by the World Bank (2022), most publications had corresponding authors based in high-income countries (78.4%). In comparison, low-income countries only had four publications with authors listed as corresponding author, reflecting 0.3% of publications and 88 from lower-middle income countries, reflecting 5.7% of publications. The remainder of the studies came from upper-middle income countries (15.6%).

Less than half of the non-review publications identified a country where the study was conducted in their abstract (39.9%). Of those, 89.5% had the same country of implementation and corresponding author. Only 27.4% of publications with authors in high-income countries mentioned the country where the study was conducted in their abstract, compared to 100% of publications from low-income countries, 76.1% from lower middle-income countries, and 64.2% of publications from upper middle-income countries. Of the 31 publications with differences between income classifications of corresponding author and where study was conducted, 30 (96.8%) had authors in high-income countries with the study conducted in upper-middle, lower-middle, or low-income countries.

There were 495 different peer-reviewed journals identified, with over three quarters publishing only one (56.0%) or two (18.3%,) relevant articles. The journal with the most publications was the Journal of Affective Disorders (23.4%) followed by the Archives of Women’s Mental Health (17.8%). Table 2 provides a bibliographic summary of top 10 authors, countries, and journals with the number of publications.

**Table 2.** Bibliographic summary of top 10 authors, countries, and journals with the number of publications.

| **Source** | **Number of Publications**<br>**(% relevant to category**<br>**total)** |
| --- | --- |
| **Authors** |  |
| Glover V | 35 (0.44%) |
| O'Connor TG | 22 (0.28%) |
| Van Den Bergh BRH | 19 (0.24%) |
| Karlsson H | 18 (0.23%) |
| Karlsson L | 18 (0.23%) |
| Reck C | 17 (0.22%) |
| Martini J | 16 (0.20%) |
| Schetter CD | 16 (0.20%) |
| Austin MP | 15 (0.19%) |
| Wittchen, HU | 15 (0.19%) |
| **Countries of Corresponding Author** |  |
| United States | 314 (20.4%) |
| United Kingdom & Northern Ireland | 141 (9.2%) |
| Australia | 136 (8.9%) |
| Canada | 124 (8.1%) |
| Iran | 76 (5.0%) |
| Turkey | 70 (4.6%) |
| Germany | 69 (4.5%) |
| Netherlands | 61 (4.0%) |
| China | 47 (3.1%) |
| Italy | 45 (2.9%) |
| **Journals** |  |
| Journal of Affective Disorders (IF: 6.5) | 115 (7.5%) |
| Archives of Women's Mental Health (IF: 4.5) | 88 (5.7%) |
| Journal of Psychosomatic Obstetrics Gynecology (IF: 3.1) | 37 (2.4%) |
| Journal Of Reproductive and Infant Psychology (IF: 3.2) | 34 (2.2%) |
| BMC Pregnancy and Childbirth (IF: 3.1) | 29 (1.9%) |
| Early Human Development (IF: 2.2) | 27 (1.8%) |
| Midwifery (IF: 2.6) | 21 (1.4%) |
| Journal of Maternal-Fetal & Neonatal Medicine (IF: 1.8) | 20 (1.3%) |
| PLOS One (IF: 3.7) | 20 (1.3%) |
| Psychoneuroendocrinology (IF: 3.9) | 19 (1.2%) |

IF = Impact Factor

In terms of the number of publications by year (Figure 1), a clear increase in publications over time was found with only 2.2% of publications dating before 1990, 3.6% between 1991-2000, 18.3% between 2001-2010, and 75.9% between 2011-2020. The year with the first publication on perinatal anxiety was 1954 by De Armond titled “A Type of Post Partum Anxiety Reaction”. The year with the largest number of publications was 2020 with 251 publications. The top 10 publications were cited a total of 5,654 times, with each being cited, on average, 39.4 times a year (Table 3).

**Figure 1.**
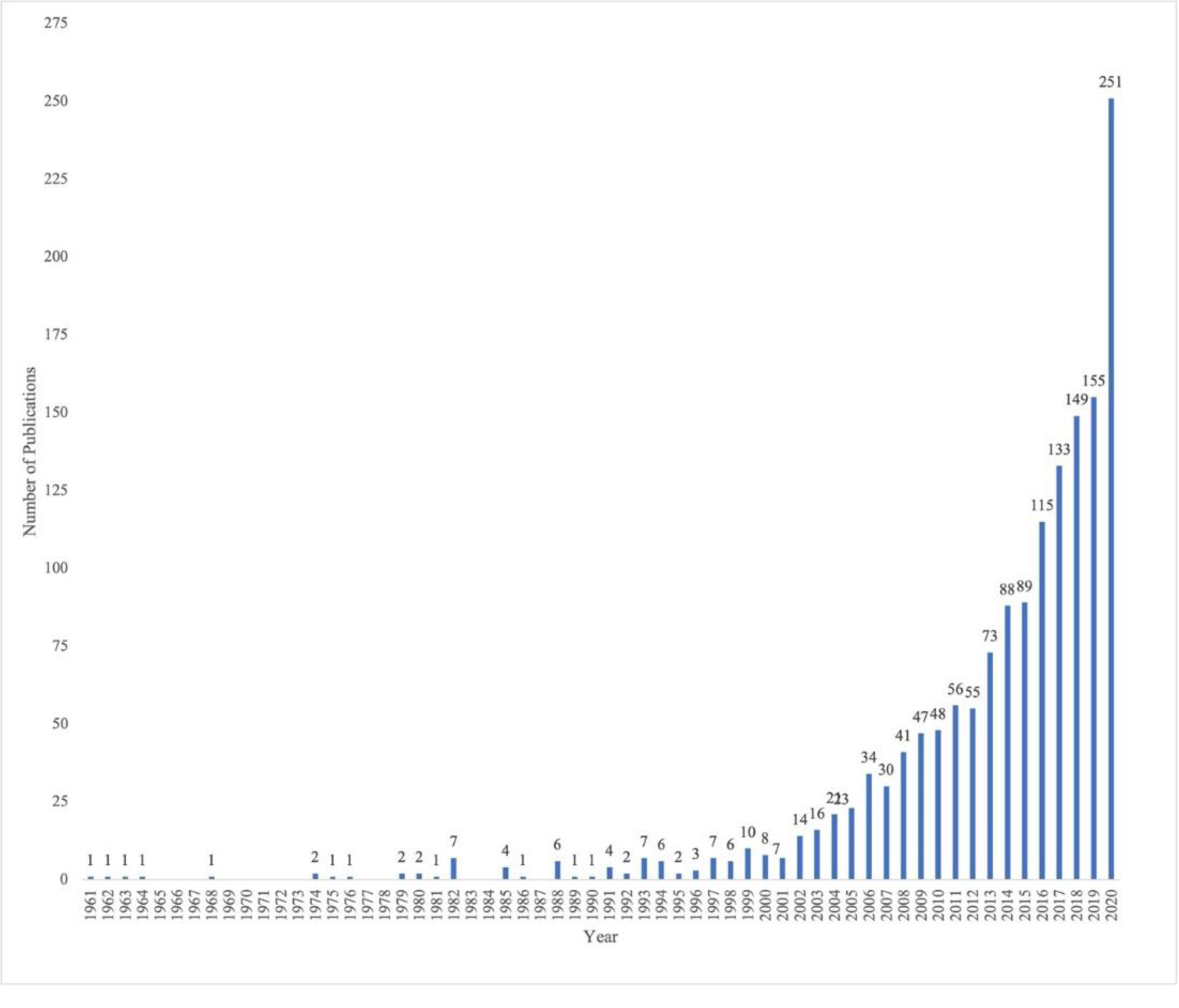
Number of publications by year.

**Table 3.** Top 10 cited publications on perinatal anxiety.

| **Title** | **Authors** | **Source Title** | **Publication Year** | **Total Citations** | **Average per Year** |
| --- | --- | --- | --- | --- | --- |
| Antenatal maternal stress and long-term effects on child neurodevelopment: how and why? | Talge, NM.; Neal, C; Glover, V | Journal of Child Psychology and Psychiatry | (2007) | 796 | 53.07 |
| The course of anxiety and depression through pregnancy and the postpartum in a community sample | Heron, J; O'Connor, TG; Evans, J; Golding, J; Glover, V | Journal of Affective Disorders | (2004) | 717 | 39.83 |
| Antenatal maternal anxiety and stress and the neurobehavioural development of the fetus and child: links and possible mechanisms. A review | Van den Bergh, BRH; Mulder, EJH; Mennes, M; Glover, V | Neuroscience and Biobehavioral Reviews | (2005) | 683 | 40.18 |
| Maternal antenatal anxiety and children's behavioural/emotional problems at 4 years - Report from the Avon Longitudinal Study of Parents and Children | O'Connor, TG; Heron, J; Golding, J; Beveridge, M; Glover, V | British Journal of Psychiatry | (2002) | 642 | 32.1 |
| Maternal stress and preterm birth | Dole, N; Savitz, DA; Hertz-Picciotto, I; Siega-Riz, AM; McMahon, MJ; Buekens, P | American Journal of Epidemiology | (2003) | 527 | 27.74 |
| Anxiety, depression and stress in pregnancy: implications for mothers, children, research, and practice | Schetter, CD; Tanner, L | Current Opinion in Psychiatry | (2012) | 513 | 51.3 |
| Depression and anxiety during pregnancy: A risk factor for obstetric, fetal and neonatal outcome? A critical review of the literature | Alder, J; Fink, N; Bitzer, Js; Hoesli, I; Holzgreve, W | Journal of Maternal-Fetal & Neonatal Medicine | (2007) | 487 | 32.47 |
| Maternal antenatal anxiety and behavioural/emotional problems in children: a test of a programming hypothesis | O'Connor, TG; Heron, J; Golding, J; Glover, V | Journal Of Child Psychology and Psychiatry and Allied Disciplines | (2003) | 441 | 23.21 |
| Identifying the women at risk of antenatal anxiety and depression: A systematic review | Biaggi, Alessandra; Conroy, Susan; Pawlby, Susan; Pariante, Carmine M. | Journal of Affective Disorders | (2016) | 434 | 72.33 |
| Stress during pregnancy is associated with developmental outcome in infancy | Huizink, AC; de Medina, PGR; Mulder, EJH; Visser, GHA; Buitelaar, JK | Journal Of Child Psychology and Psychiatry and Allied Disciplines | (2003) | 414 | 21.79 |

### Visualization Mapping

Visualization mapping was conducted with the full list of relevant publications (n=2,203) to reflect the breadth of relations on perinatal anxiety. Therefore, this analysis included all publications related to perinatal anxiety (e.g., as a primary, secondary, and covariate outcome), not just those with it as a primary focus.

### Co-authorship

Co-authorship was defined as the number of times two authors were listed together in an author list. Of the 7,712 authors, 197 authors were identified that had a minimum of 5 citations each, reducing authors first name to initial only. Removing non-connected authors, only 118 authors remained that were connected to each other (Figure 2). The authors with the highest number of citations were Glover V (n=7,108 with 41 documents), Heron, J (n=2,742 with 8 documents), and O’Connor, T (n=2,638 with 6 documents). The authors with the highest number of links, or connections to another author, were Karlsson, H (n=69), Karlsson L (n=69), and Glover V (n=66).

**Figure 2.**
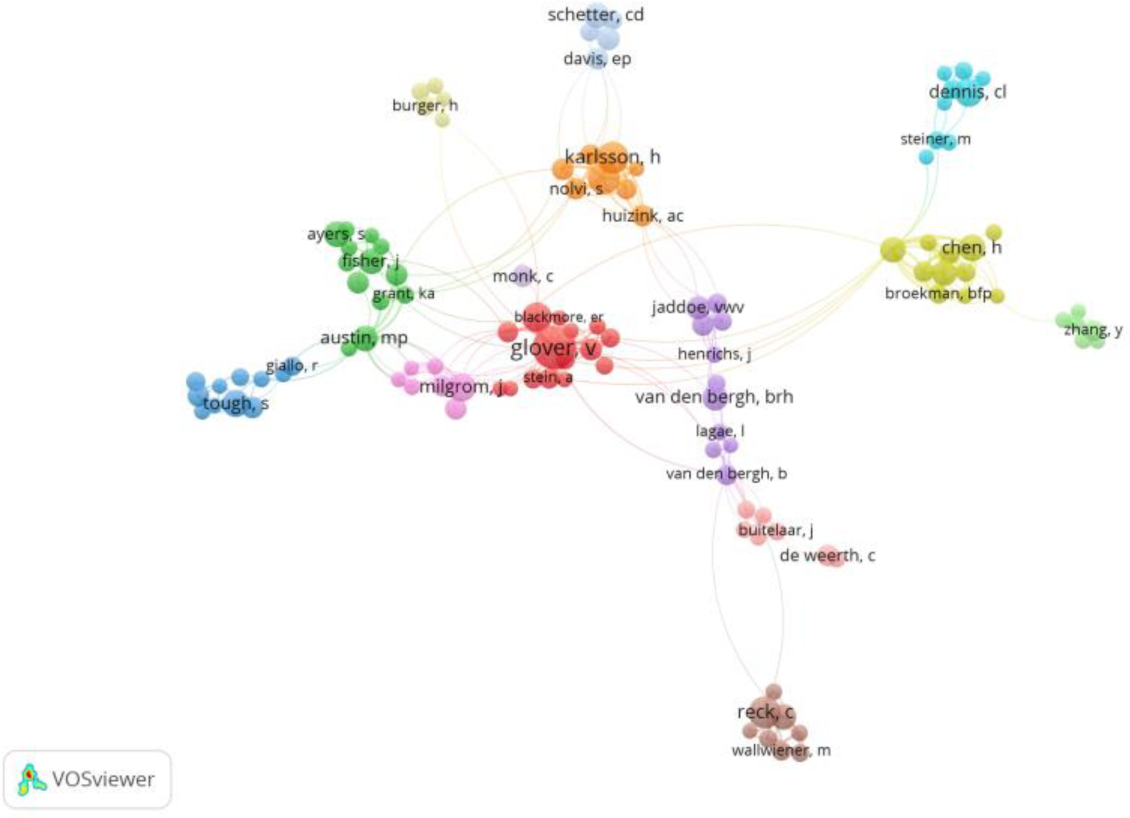
Network visualization of co-authorship by author with at least 5 publications, including only connected authors (n=118)

Next, we visualized the network between co-authored publications by countries of corresponding author that had at least one citation and one publication. Of the 79 countries, 76 met this inclusion criteria. Excluding five countries which had no connection to each other, a total of 71 countries were visualized in Figure 3. This reflects the degree of co-authorship between countries of corresponding authors (shown through the links) as well as most frequent countries publishing on this topic (shown through the nodes). The countries with the highest number of citations were the United States (n=26,368 with 581 documents and 273 link strength), followed by England (United Kingdom; n=15,716 with 266 documents and 248 link strength) and Canada (n=9,368 with 225 documents and 148 link strength).

**Figure 3.**
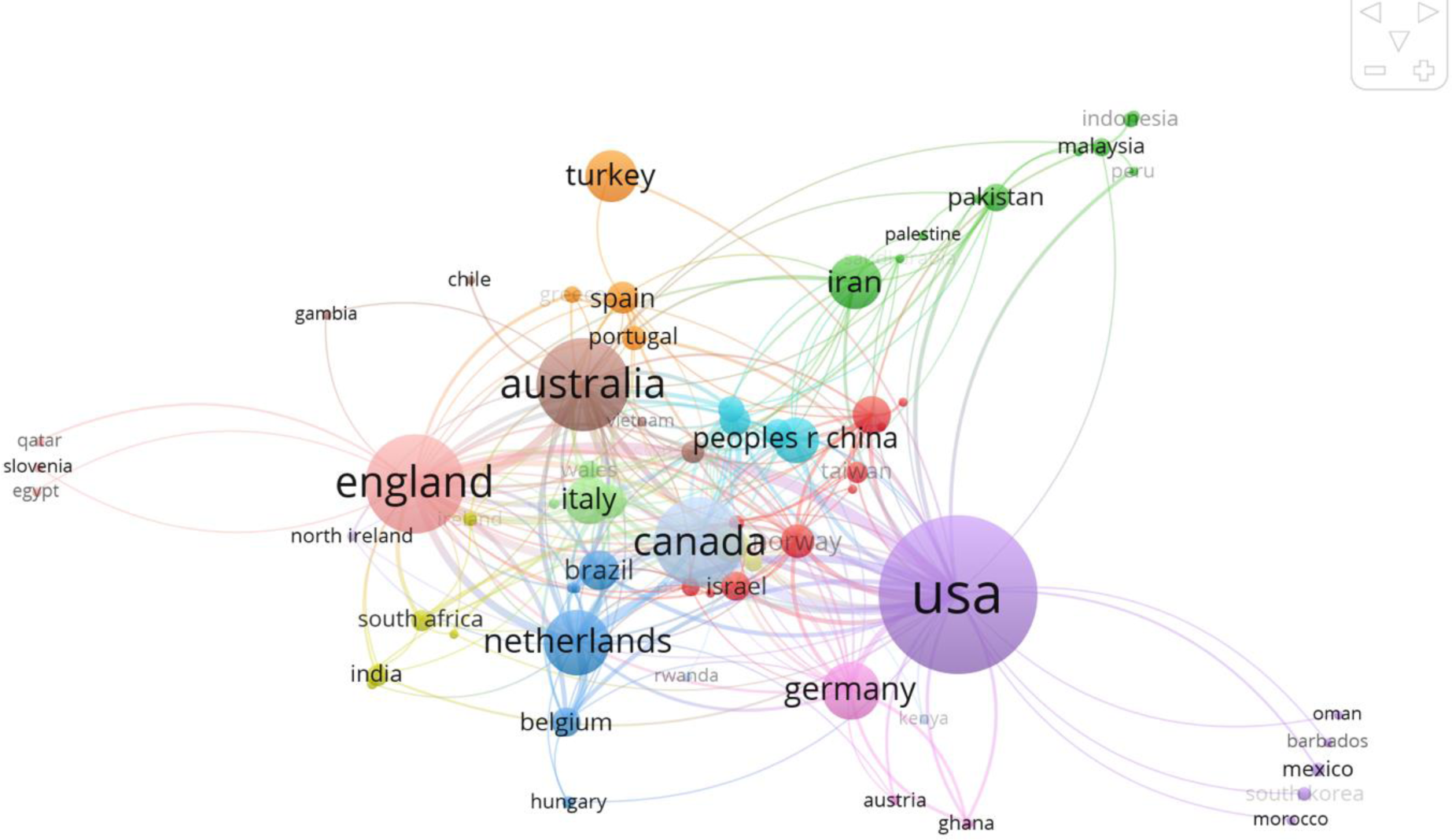
Network visualization of co-authorship between 71 countries with at least one publication and one citation

### Keyword co-occurrence

Keywords are selected by the authors to describe the publication, which are used by the journals to tag for relevant themes. In the network visualization of all keywords used by the authors which appeared a minimum of 10 times, 297 keywords met the threshold from 4,916 possible keywords. Reducing the network visualization of all keywords which appeared a minimum of 50 times, 53 keywords met the threshold (Figure 4). This visualization illustrates the relationship between keywords and how often they are used with other keywords. The keywords with the highest number of occurrences were anxiety (n=916), pregnancy (n=856), and depression (n=688).

**Figure 4.**
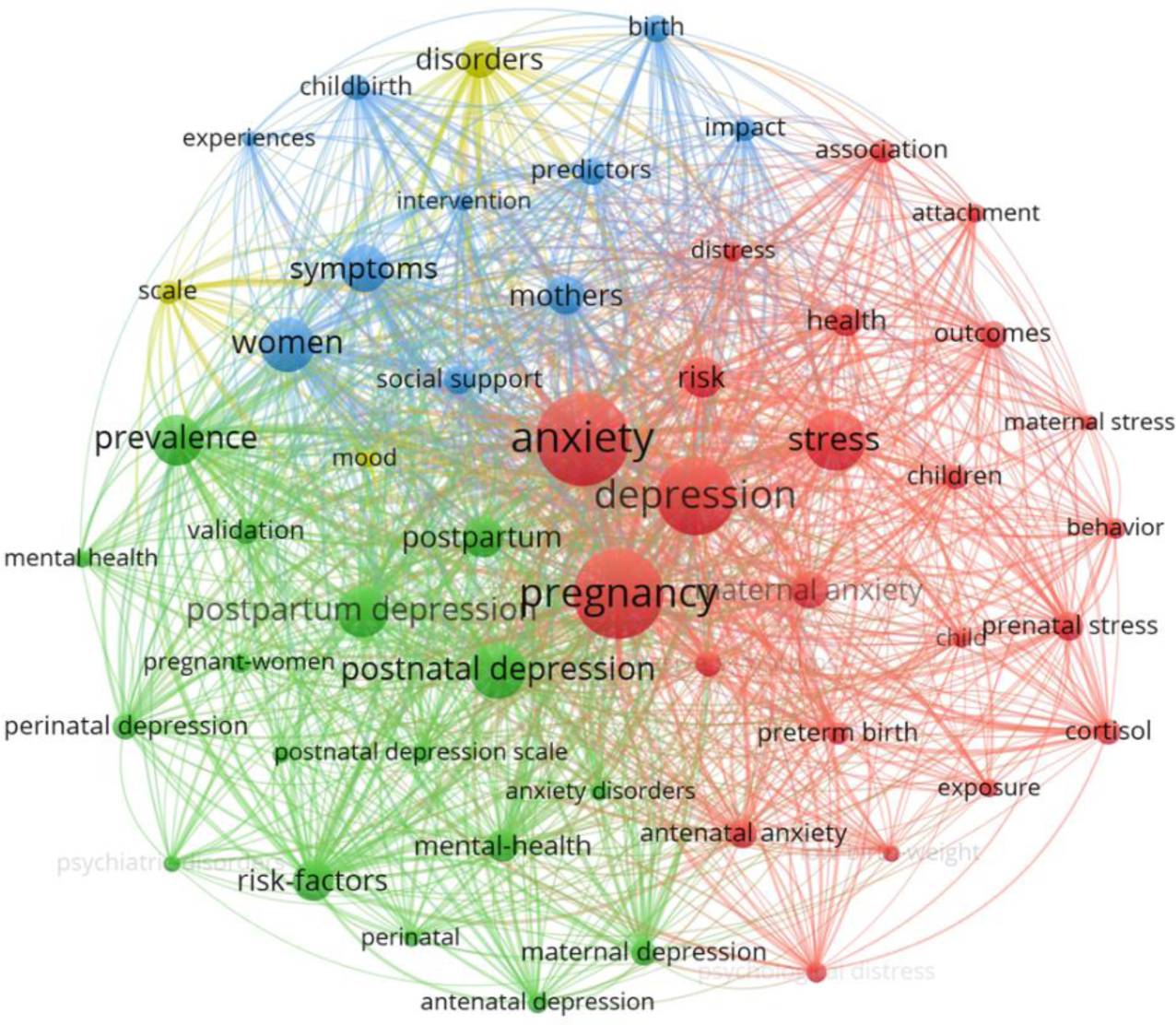
Network visualization of keyword co-occurrence which appeared at least 50 times (53 keywords)

We also ran a keyword term analysis which used all the words in the title and abstract to identify the most frequently used terms. Using a binary approach, where the presence or absence was counted, regardless of frequency, the VOSviewer program was set to identify terms that appeared at least 10 times or 50 times, excluding structured abstract labels and copyright statements. Of the total 33,314 terms, there was 1,318 terms that appeared at least 10 times and 243 terms that appeared at least 50 times. The top terms here were anxiety (n=1,534), study (n=1,307), and pregnancy (n=1,202).

### Citation, Bibliographic Coupling & Co-Citation

We analyzed the number of citations which indicated relatedness based on the number of times one publication cited another. Including all publications with at least one citation, 2,076 publications were included. The publications with the largest number of links to other publications were Heron (2004) with 277 links, Dennis (2017) with 183 links, and Van Den Bergh (2005) with 180 links. The publications with the larger number of citations were Talge (2007) with 828 citations, Heron (2004) with 745 citations, and Van Den Bergh (2005) with 711 citations.

Next, we conducted a bibliographic coupling analysis to calculate the number of shared references between publications (i.e., two publications that both cite another publication) with at least one citation (n=2,076) and 50 citations (n=427). We also examined the co-citation for the number of times publications, journals, and first authors were cited together in a third publication’s citations. Of the 51,334 cited references, 1,249 were cited at least 10 times and 107 were cited 50 times. The publications with the greatest number of citations were Cox (1987) (n=625), O’Connor (2002) (n=282) and Spielberger (1983) (n=202). We also analyzed the co-citation for first authors, which means two first authors are both cited in the same publication. Among the 32,510 different first authors, 1,587 were cited together at least 10 times and 197 at least 50 times. The authors with the greatest number of citations were Cox JL (n=730), Field T (n=635), and O’Connor TG (n=542).

Of the 12,395 different journals, Figure 5 illustrates the co-citation for journals cited at least 10 times (n=1,019) This represents co-citations between journals, with the size of the node representing the number of publications at that journal. Additionally, the distance between different nodes shows a relationship in that the smaller the distance, the higher the citation frequency. The sources with the greatest number of citations were Journal of Affective Disorders (n=3,892), Archives of Women’s Mental Health (n=2,681) and British Journal of Psychiatry (n=1,993).

**Figure 5.**
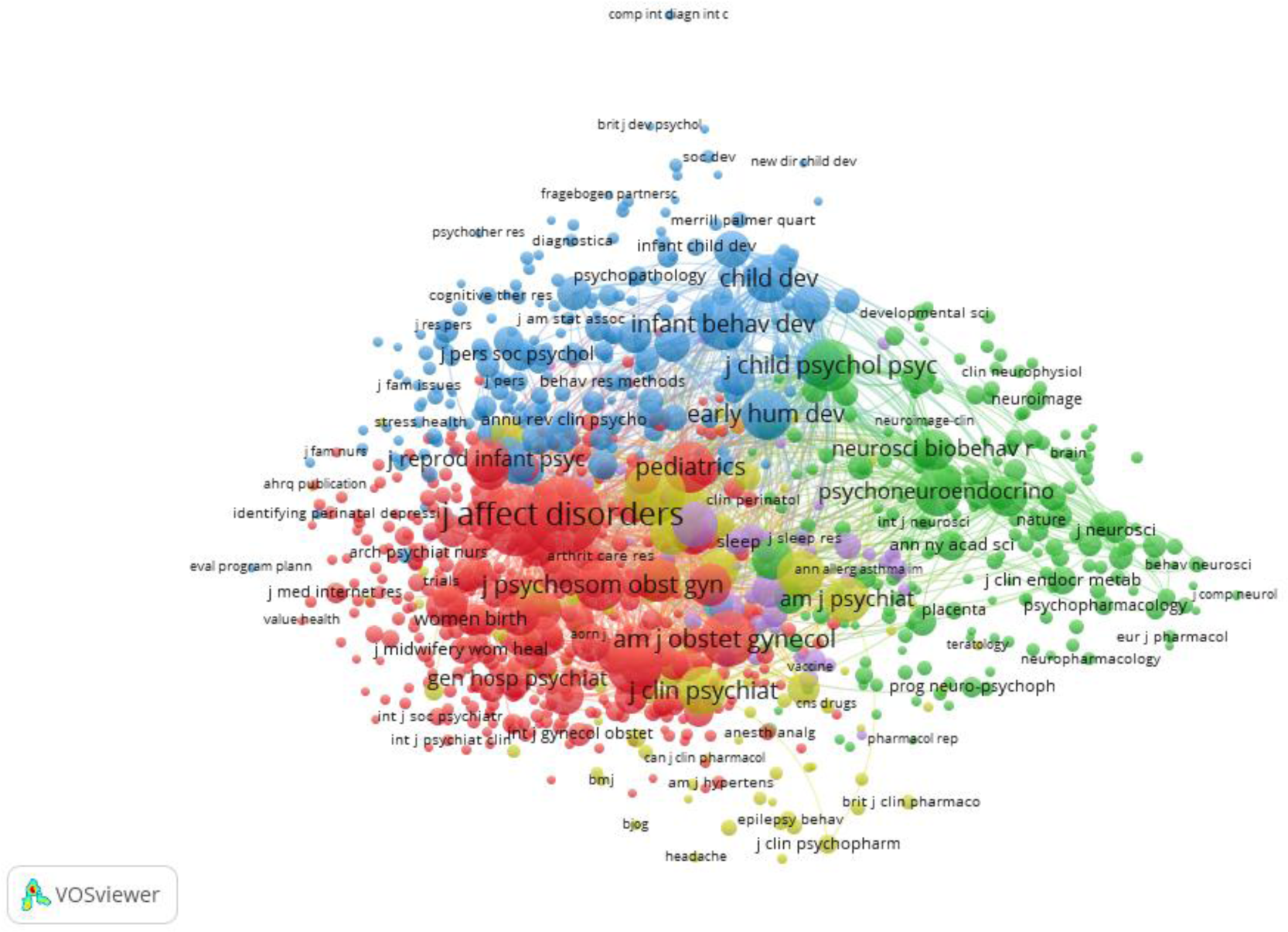
Co-citation between journals of publications with at least 10 citations (n=1,019)

## Discussion

This is the first study to map the literature on anxiety across the perinatal period from pregnancy to the first year postpartum with publications between 1920 and 2020. We found that there has been a dramatic increase in the number of publications on perinatal anxiety over time since the first publication in 1954. While publications have been circulated in 495 different peer-reviewed journals, over half (56.0%) have only published one related article. Most publications focused on antenatal anxiety and maternal anxiety, suggesting that postpartum anxiety, intrapartum anxiety, and paternal anxiety are understudied. There is also a significant geographical divide with most corresponding authors based in high-income countries.

Between 2011 and 2020, almost three-quarters of publications related to perinatal anxiety were published, indicating a high level of interest in this field of research and may reflect new funding related to mental health research. Interestingly, all but two of the top 20 cited publications were published prior to 2010. This suggests that despite a significant growth in publications, there are seminal papers that are foundational for the field. Additionally, publications have been published in a wide variety of academic journals, with considerable interdisciplinary diversity in authorship and focus ranging from psychiatry to nursing to behavioral sciences. While there are some journals that published more frequently on this topic, over half of the journals identified cited only one relevant publication, suggesting a lack of a primary journal dedicated to this topic. This resulted in some potential difficulty in accessing related work yet could also suggest that perinatal anxiety is increasingly being recognized as an important issue across varied health disciplines. Not having key journals may limit the field in that it could limit the impact of the research if others cannot find it. The top two journals with the most publications (Journal of Affective Disorders and Archives of Women’s Mental Health) had the highest impact factors (IF), which are used to measure the number of times selected articles are cited within the last few years, with higher IF indicated a higher rank of journal. It will be important to see if, over time, a narrowing of publications among certain journals emerges or if publications continue to be published in a wide variety of journals.

An interesting aspect that emerged during this bibliometric review was the amount non-relevant publications related to perinatal anxiety that resulted in the need to screen all included studies. This illustrates the challenges with citation analysis in that studies that use key terms (in this case, perinatal anxiety) may be captured but anxiety may not be a primary focus. This is evident in the fact that only 69.6% of publications that were related to perinatal anxiety had it as a focus of the paper, rather than a co-reported or covariate outcome. Further, over half of the studies co-reported on perinatal depression. This is not necessarily a surprise as depression in the perinatal period has been more widely studied and there is established correlations between depression and anxiety (Falah-Hassani et al., 2016; Littleton et al., 2007). The co-relation between depression and anxiety is also re-affirmed in the keyword and term analysis, with depression being a top keyword. This suggests that these terms are co-related in the literature, often appearing together. This suggests that the field of perinatal anxiety publication often researched in relation to other mental health and perinatal outcomes.

An additional gap in the current literature is the lack of focus on non-maternal perinatal anxiety, with only 6.0% of publications reporting on paternal perinatal anxiety. There is a significant gap in the available evidence on the perinatal anxiety experience of the non-birthing partner, despite one in five fathers experiencing perinatal anxiety (Leach et al., 2016; Leiferman et al., 2021). As the field continues to evolve, it is likely that shifts will occur in terms of focusing where the gaps are, such as the emerging trend of using more inclusive terminology around birthing people (Bamberger and Farrow, 2021; Crossan et al., 2023) and the inclusion of perinatal anxiety outcomes for all parents.

In terms of geographical influence, the countries that dominate publications are from primarily high-income countries, including United States, England, and Australia as the top three. This was also reflected in the visualization of co-authorship between countries, where there are weaker links between low-income and lower-middle income countries compared to links between other high-income countries. This suggests that there is a dearth of publications in perinatal anxiety from low-and lower-middle income countries (LMICs), which was supported by the finding that only four studies were from low-income countries. The scarcity and lack of mental health research in LMICs broadly has been identified previously by others, which has been linked to both lack of publication in indexed journals as well as a lack of human resources and experts in this research field (Alloh et al., 2018; Razzouk et al., 2010). This trend may also be related to fewer funding opportunities and the greater stigma related to mental health in LMIC (Alderdice and Newham, 2016; Alloh et al., 2018; Razzouk et al., 2010). This discrepancy is not associated with a lower perinatal anxiety rate, as Dennis and colleagues (2017) found in their systematic review that the perinatal anxiety rate was actually higher in LMICs than high-income countries. Thus, this is a significant gap in the current literature as publications are predominantly from high-income countries yet LMICs have higher rates of perinatal anxiety. It is important that future work not only focuses on perinatal anxiety in LMICs but also researchers from LMICs are engaged as co-authors in the research to enhance representation from LMICs. Furthermore, it is important that work that is done in low and lower-middle income countries have authors from those countries listed as corresponding authors, rather than from high-income countries which was predominant in this analysis.

In terms of design, only 1.5% of studies were qualitative, with the majority being either quantitative design or reviews. This is a gap in the literature as qualitative research can provide insight that quantitative designs cannot, including the ability to discover reasons for observed patterns, especially the invisible or non-anticipated ones (Busetto et al., 2020). Qualitative research on perinatal anxiety can help explain the ‘why’ behind the prevalence, the ‘what’ behind the symptoms, and the ‘how’ and ‘who’ for effective intervention implementation.

There was also a significant number of authors identified, with the top three being: Glover V, O’Connor TG, and Van Den Bergh BFH, in terms of number of total publications as well as mapping co-authorship, citations, and bibliographic coupling. This suggests that the work of these authors is inter-related, highly cited, and collaborative. Unlike other specialities (Ellis et al., 2019), there is considerable interaction with other authors, suggesting a diverse, emerging field with large variation in authors and ideas.

## Limitations

While this study has several strengths, there are some limitations that much be acknowledged. Due to the data extraction and analysis approach used, we only used Web of Science to identify relevant articles. While this database did yield the largest number of citations, it likely did not capture all studies, thus this bibliometric review may be under-reporting the number of publications on perinatal anxiety. Another limitation was that due to the number of publications identified not related to perinatal anxiety, manual screening of each publication was required to ensure it was related to perinatal anxiety. While double verification was completed, there was some possibility for excluding eligible publication if an abstract was not clearly in reporting perinatal anxiety as full texts were not reviewed. Finally, researchers who publish differently with their first and/or second initial (Smith B and Smith BA) may have ended up with different outcomes in the visualization and the bibliometric analysis, which may have resulted in either an over or under estimation. For the bibliometric analysis, authors were collapsed when possible (e.g., Smith B and Smith BA were collapsed) but were not when it was unclear (e.g., Smith BA and Smith BC were not collapsed). For the visualization, VOSviewer collapses by first initial so regardless, Smith B would include Smith B, Smith BA, and Smith BC.

## Conclusion

Perinatal anxiety as a field of study has been increasing exponentially and will likely continue to be a prevalent focus in the future. Often published in relation to depression, perinatal anxiety is a growing field with consistent authors identified as experts in the field. Future work related to non-birthing parent perinatal anxiety, qualitative or mixed method approaches, and postpartum and intrapartum only foci may be warranted. Continued work in this field is warranted to further understand the unique and complex relationships that perinatal anxiety has on a variety of outcomes for birthing persons, their partners, and their infants.

## Data Availability

The data that support the findings of this study are available from the corresponding author upon reasonable request.

## Acknowledgements

The authors would like to acknowledge the contributions of Brendan Pugh from Dalhousie University for assistance with data extraction.

